# Spread of dual-class drug-resistant of *Mycoplasma genitalium* in Tokyo, Japan, 2023–2025

**DOI:** 10.1101/2025.09.10.25335488

**Authors:** Ryuha Omachi, Kazuo Imai, Akihiro Sato, Masashi Tanaka, Hitomi Mizushina, Keita Takeuchi, Nobuaki Mori, Takuya Maeda

## Abstract

**Background:** The increasing prevalence of *Mycoplasma genitalium* (MG) strains harboring macrolide and fluoroquinolone resistance-associated mutations (MRMs and QRMs, respectively) is a growing global concern. However, data on resistance patterns and genetic diversity in Japan remain limited.

**Objectives:** This study investigated MRMs and QRMs, genetic diversity using *mgpB* and *MG309* typing, and their association with treatment outcome in MG strains collected in Tokyo, Japan, between 2023 and 2025.

**Patients and methods:** Between 2023 and 2025, 188 clinical samples from 162 MG-positive patients were analyzed. Resistance mutations in *23S rRNA, parC*, and *gyrA* were sequenced, and molecular typing was performed. Treatment outcomes were assessed using test-of-cure results.

**Results:** MRMs and QRMs in *parC* S83I and *gyrA* were identified in 94.4%, 65.5%, and 22.5% of samples, respectively. Dual-class resistance (MRMs + QRMs) was found in 89.4% of strains.

Phylogenetic analysis based on *mgpB* and *MG309* typing revealed the emergence of dual-class drug-resistant clonal complexes, particularly those harboring *mgpB* alleles 79, 140, 161, and 184. Dual-QRMs were significantly associated with quinolone treatment failure (52.4% vs. 23.5%, p = 0.016).

**Conclusions:** Dual-class drug-resistant MG strains, including emerging clonal complexes, are spreading in Tokyo. These findings emphasize the need for continued molecular surveillance and prudent antimicrobial use to preserve treatment efficacy.

## Main text

*Mycoplasma genitalium* (MG) is a sexually transmitted infection that can infect the urethra, cervix, and rectum (1). In Japan, urethral MG has been detected in 22% of men with symptomatic non-gonococcal urethritis (2). Macrolides, fluoroquinolones, and tetracyclines are commonly used to treat MG infections; however, the increasing prevalence of macrolide and quinolone dual-class drug-resistant strains has become a growing public health concern (3).

Macrolide resistance-associated mutations (MRMs) of MG have been identified as single nucleotide mutations at positions A2058 and A2059 in the V region of the *23S rRNA* gene (*Escherichia coli* numbering) (4). These MRMs induce structural changes in the ribosomal antibiotic binding site, thereby reducing the effectiveness of macrolide antibiotics (4). Quinolone resistance-associated mutations (QRMs) are linked to mutations in the quinolone resistance-determining regions of the *parC* and *gyrA* genes, which encode DNA topoisomerase IV and gyrase, respectively (5). The amino acid changes in the quinolone resistance-determining regions of *parC* and *gyrA* are associated with an increase in the minimum inhibitory concentration of moxifloxacin and sitafloxacin (5). Especially, amino acid changes in ParC (e.g., S83I) and GyrA (e.g., G93, M95, and D99) are associated with moxifloxacin and sitafloxacin treatment failure (6-8). Although high antimicrobial resistance rates of MG have been reported among the men who have sex with men (MSM) population in Tokyo (9, 10), data on resistance in the non-MSM population in Japan remain limited (11).

Evaluating the diversity of drug-resistant strains through molecular typing methods is essential for identifying high-risk populations, estimating transmission routes, and formulating effective intervention strategies. Sequence typing methods such as *mgpB* (MG191) adhesin gene single-nucleotide polymorphism typing (12) and *MG309* short tandem repeat typing (13) have shown high discriminatory power (14) and have been widely used for molecular surveillance globally (15-17). However, in Japan, the evaluation of MG strain diversity, including drug-resistant strains, using *mgpB* and/or *MG309* typing has not been conducted sufficiently to date (18, 19). To optimize the effectiveness of treatment and prevent the further spread of drug-resistant MG strains, it is important to continuously monitor the prevalence and diversity of resistance-associated mutations.

This study investigated the prevalence and diversity of MRMs and QRMs in MG as well as their association with treatment outcome in the clinical setting in Tokyo, Japan.

## Materials and Methods

### Patients and sample collection

Adult patients (≥18 years old) in whom MG infection was diagnosed at KARADA Internal Medicine Clinics (Shibuya and Gotanda, Tokyo, Japan) between 2023 and 2025 were enrolled in this study. MG was detected by cobas MG/TV (Roche, Basel, Switzerland), multiplex PCR for STD Mycoplasma Nucleic Acid Testing (LSI Medience Corporation, Tokyo, Japan), and laboratory-developed quantitative PCR testing of the *MgPa* gene of MG (20). Residual first-void urine and vaginal swab samples after routine testing were utilized in this study. The samples were stored at −80°C until DNA extraction. Patient information was collected retrospectively from the hospital electronic medical records. This study was reviewed and approved by Saitama Medical University Hospital (approval number: 2023-026). Informed consent was obtained by opt-out via the hospital’s website.

### DNA extraction

First-void urine and suspension of vaginal swabs (1 mL) were centrifuged at 15,000 × *g* for 15 min at room temperature, and the sediment was resuspended in 140 μL phosphate-buffered saline. DNA was extracted from 140 μL of the suspension, using a QIAamp Viral RNA Mini Kit (Qiagen, Hilden, Germany), and eluted in 60 μL of 10 mM Tris-HCl buffer, pH 8.5.

### Sequencing of drug resistance-associated genes and *mgpB*-*MG309* typing

*23S rRNA, gyrA, parC, mgpB*, and *MG309* were analyzed in this study. Nested PCR amplification of the genes (12, 13, 21, 22) was performed using KOD One PCR Master Mix (Toyobo, Tokyo, Japan). The primer sequences are shown in **Supplementary Table S1**. The amplicons were analyzed by 1% agarose gel electrophoresis with ethidium bromide staining, and the PCR products were purified using an ExoSAP-IT (Applied Biosystems, Waltham, MA) or QIAquick PCR Purification Kit (Qiagen). Direct Sanger sequencing was performed by Eurofins Genomics (Tokyo, Japan).

### Sequencing analysis

The obtained sequence data of *23S rRNA, gyrA*, and *parC* were compared with the corresponding regions of the MG reference strain G37 (GenBank: L43967.2) to identify mutations. The sequence types (STs) from *mgpB*-*MG309* typing were determined via the PubMLST BIGSdb database (https://pubmlst.org/organisms/mycoplasma-genitalium) (23). The phylogenetic tree of the concatenated *mgpB* and *MG309* sequence was constructed using IQtree ver. 2.4 (24). Phylogenetic trees were created using iTOL (https://itol.embl.de/).

### Statistical analysis

Continuous variables with a normal distribution are expressed as the mean (±standard deviation) and those with a non-normal distribution as the median (interquartile range) and were compared using Student’s *t*-test or the Mann–Whitney *U* test, as appropriate. Categorical variables are presented as frequency and percentage (%) and were compared using a chi-squared test or Fisher’s exact test, as appropriate. A two-sided *p*-value of <0.05 was considered statistically significant. All statistical analyses was conducted using R ver. 4.1.2 (R Foundation for Statistical Computing, Vienna, Austria).

## Results

### Baseline characteristics of the participants

During the study period, 162 patients with confirmed MG infections were enrolled, and a total of 188 clinical samples (152 first-void urine and 36 vaginal swabs) were collected and subsequently submitted for laboratory analysis. The baseline characteristics of the study participants are shown in **Table 1**. Among the 162 patients, the median age was 30 years, with an interquartile range of 25–39 years, and 79.0% (*n* = 128) of the patients were male. Regarding potential risk factors for infection, 33 patients (20.4%) reported engaging in sexual activity with commercial sex workers. Thirteen patients (8.0%) reported working as commercial sex workers. Sexual activity with anonymous partners was reported by 50 patients (30.9%), while 74 patients (45.7%) reported having sexual activity with a regular partner. The number of respondents who did not disclose information for each category was 29 (17.9%), 29 (17.9%), 51 (31.5%), and 51 (31.5%), respectively.

**Table 1.** Clinical characteristics of the study participants.

|  | All patients<br>(*n* = 162) |
| --- | --- |
| Age, years, median [interquartile range] | 30 [25–39] |
| Male, *n* (%) | 128 (79.0) |
| Female, *n* (%) | 34 (21.0) |
| **Risk of infection, *n* (%)** |  |
| Use of commercial sex workers | 33 (20.4) |
| Unknown | 29 (17.9) |
| Working as a commercial sex worker | 13 (8.0) |
| Unknown | 29 (17.9) |
| Sexual activity with anonymous partners | 50 (30.9) |
| Unknown | 51 (31.5) |
| Sexual activity with a regular partner | 74 (45.7) |
| Unknown | 51 (31.5) |
| **Initial treatment, *n* (%)** |  |
| Minocycline 7 days + sitafloxacin 7 days (sequential) | 137 (84.6) |
| Moxifloxacin 10 days | 5 (3.1) |
| Sitafloxacin 7 days | 2 (1.2) |
| Minocycline 7 days | 5 (3.1) |
| Doxycycline 7 days | 2 (1.2) |
| Other | 3 (1.9) |
| None | 8 (4.9) |
| **Initial treatment outcome, *n* (%)** |  |
| Cure | 61 (37.7) |
| Failure | 33 (20.4) |
| Unknown | 68 (41.9) |

The most common initial treatment regimen in 137 patients (84.6%) was the sequential administration of minocycline for 7 days followed by sitafloxacin for 7 days. Other regimens included moxifloxacin for 10 days (*n* = 5, 3.1%), sitafloxacin for 7 days (*n* = 2, 1.2%), minocycline for 7 days (*n* = 5, 3.1%), and doxycycline for 7 days (*n* = 2, 1.2%). Eight patients (4.9%) did not receive any initial treatment. Treatment outcome was assessed based on the results of a test of cure (TOC) conducted between 28 and 90 days following the initiation of treatment. Outcome data were available for 94 patients (58.0%), among whom 61 (64.9%) achieved microbiological cure, while 33 (35.1%) experienced treatment failure. The remaining 68 patients (42.0%) did not have TOC results available.

Among the 33 patients who experienced initial treatment failure, 31 received additional treatment, and 23 of them finally achieved microbiological cure (**Supplementary Table S2**). The number of treatment cycles required for cure ranged from 2 to 7. Eight patients were cured with the sequential administration of minocycline for 7 days followed by sitafloxacin for 7 days, and 10 patients were cured through long-term minocycline or doxycycline therapy for 14 to 28 days. Three patients received a combination of moxifloxacin for 7 days and metronidazole for 7 days, and another patient was treated with spectinomycin (2 g/day) for 7 days and moxifloxacin for 10 days.

### Analysis of resistance-associated genes

Of the 188 clinical samples, *23S rRNA, parC*, and *gyrA* were successfully sequenced in 160 samples collected from 142 patients. The results of sequence analysis for the 142 clinical samples collected prior to initial treatment are summarized in **Table 2**. Among the 142 samples, mutations in *23S rRNA* were highly prevalent (*n* = 134, 94.4%). The most common mutation was A2059G (*E. coli* numbering), found in 94 samples (66.2%), followed by A2058G in 29 samples (20.4%) and A2058T in 11 samples (7.7%). Wild-type (WT) sequences were observed in only 8 samples (5.6%). Mutations in *parC* were also detected frequently in 133 samples (93.7%), particularly at codons 83 and 87. The S83I (G248T) mutation was the most common (*n* = 93, 65.5%). Other mutations included S83N (G248A, *n* = 20, 14.1%), D87N (G259A, *n* = 7, 4.9%), D87Y (G259T, *n* = 6, 4.2%), and several others occurring at lower frequencies. The WT sequence was found in 9 samples (6.3%). In *gyrA*, the WT sequence was dominant, identified in 110 samples (77.5%). Mutations resulting in amino acid changes were observed less frequently in 32 samples (22.5%), with M95I (G285A/T) in 20 samples (14.1%), M95V (A283G) in 5 samples (3.5%), D99N (G295A) in 5 samples (3.5%), and G93C (G277T) in 2 samples (1.4%).

**Table 2.**
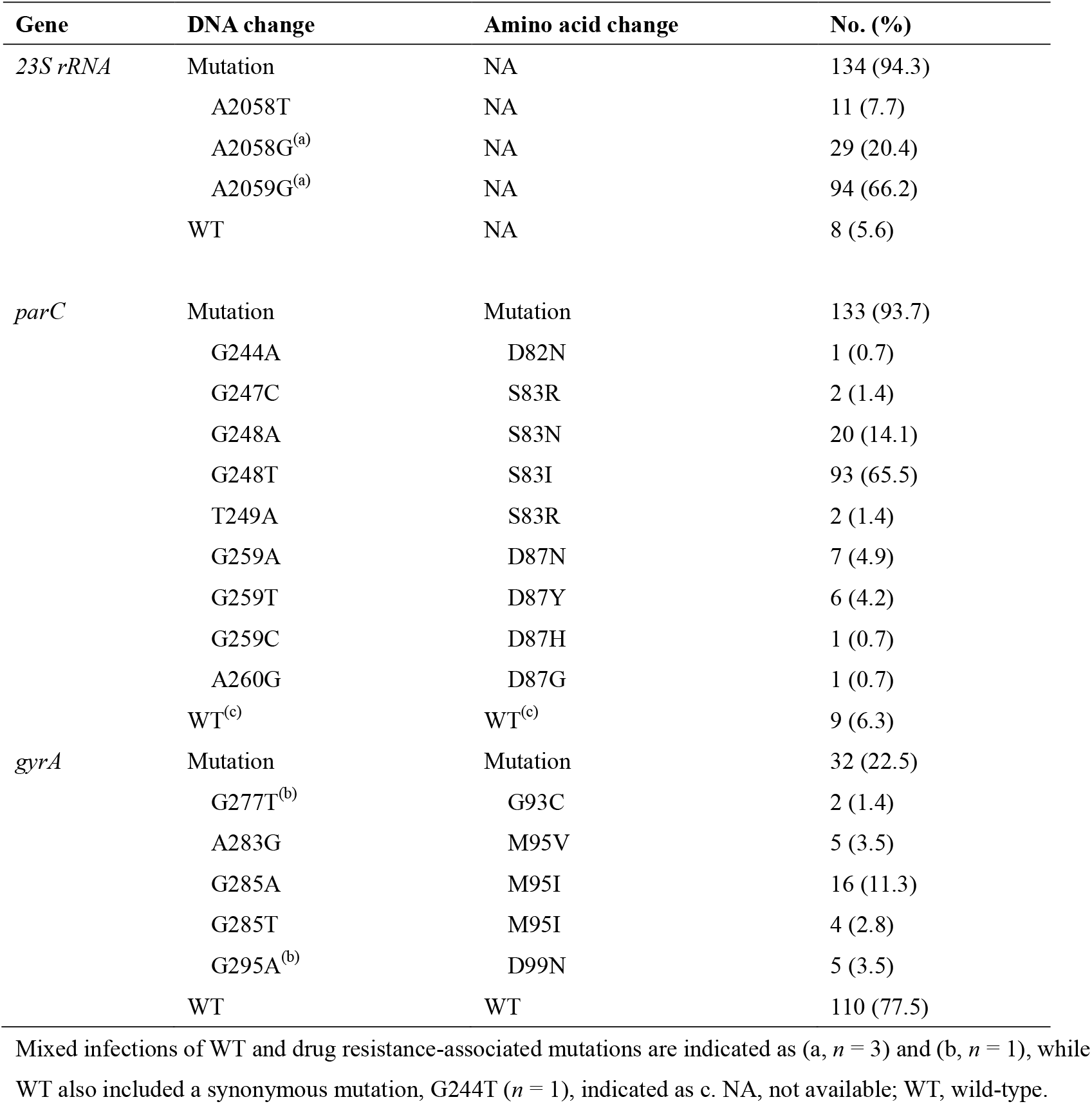
Frequency of mutations in drug resistance-associated genes in 142 samples.

### Analysis of dual-class drug resistance

Among the 142 samples, dual-class mutations in both *23S rRNA* and *parC*—MRMs and QRMs, respectively—were frequently observed (*n* = 127, 89.4%) (**Supplementary Table S3**). Dual-QRMs, characterized by mutations in both *parC* and *gyrA*, were found in 32 samples (22.5%), with only 2 exceptions in which dual-class MRMs and QRMs were present. The most common genotype combination—A2059G in *23S rRNA* and S83I in *parC*—was found in 82 samples (57.7%), with 26 of these (18.3%) also carrying *gyrA* mutations. Although the overall proportion was small, mutations in *parC* (S83R and D87Y/N) were also associated with mutations in *gyrA*.

### *mgpB*-*MG309* typing and phylogenetic analysis

Of the 160 samples that were sequenced for *23S rRNA, parC*, and *gyrA*, 135 were fully typed at *mgpB*-*MG309*. Sequential samples from the same patient with identical STs were considered duplicates and excluded, resulting in 126 unique samples collected from 121 patients for analysis. Typing identified 27 distinct *mgpB* alleles, including 12 novel alleles (alleles 387–398), and 23 distinct *MG309* alleles, with one novel allele (allele 136). Combined *mgpB*-*MG309* typing revealed 61 distinct STs, including 41 novel STs (ST561, 581, 585, 588, 596, 600, 605–631, 633, and 635– 640) (**Supplementary Table S4**).

Phylogenetic analysis of concatenated *mgpB* and *MG309* sequences grouped the strains into two distinct clusters, A and B, with 95% ultrafast bootstrap support based on IQ-TREE analysis (**Figure 1**). Cluster A (*n* = 27) was composed primarily of strains harboring *mgpB* alleles 2 (*n* = 7), 140 (*n* = 4), and 79 (*n* = 8). The most frequent STs within this cluster were ST1 (*mgpB*-*MG309* allele 2-3, *n* = 3) and ST209 (79-4, *n* = 3). All strains carrying *mgpB* alleles 140 and 79 exhibited dual-class resistance mutations of MRMs and *parC* S83I. The majority of strains analyzed in this study belonged to Cluster B (*n* = 99). The dominant *mgpB* alleles in this cluster were 7 (*n* = 28), 161 (*n* = 38), and 184 (*n* = 10). The most frequent ST was ST78 (7-4; *n* = 11). Except for 3 strains, all strains with *mgpB* alleles 161 and 184 harbored MRMs and the *parC* S83I mutation. Specifically, ST368 (161-3, *n* = 4), ST441 (161-4, *n* = 7), ST199 (161-6, *n* = 7), ST596 (161-8, *n* = 4), and ST622 (184-4, *n* = 3) were associated with a 100% prevalence of the *23S rRNA* A2059G and *parC* S83I mutations. Mutations in *gyrA* were sporadically detected across various *mgpB* alleles. No associations were observed between phylogenetic clustering and potential risk factors for infection.

**Figure 1.**
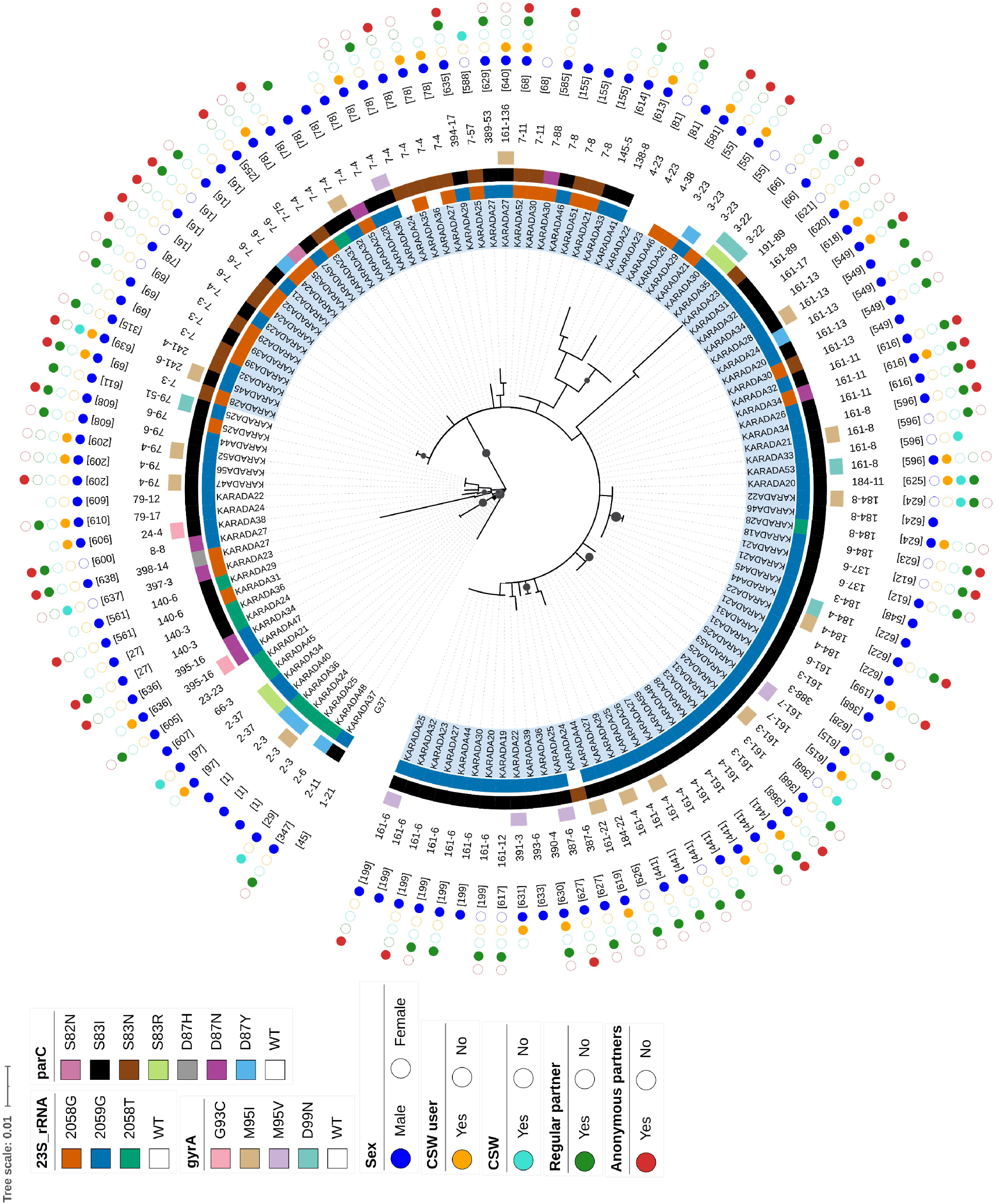
Phylogenetic tree of concatenated *mgpB* and *MG309* sequences. The phylogenetic tree was constructed using the maximum likelihood method based on 126 clinical samples. The G37 strain (GenBank: L43967.2) was used as a reference. Cluster B is highlighted in blue. From the inner to the outer rings, the plot displays the following information: *23S rRNA, parC, gyrA*, and *mgpB*– *MG309* alleles, sequence type (ST), sex, and risk of infection (including use of commercial sex workers [CSW], working as a CSW, and sexual activity with regular or anonymous partners). Black circles on nodes represent >95% ultrafast bootstrap support. The scale bar indicates a branch length corresponding to a genetic distance of 0.01.

### Association between QRMs and treatment outcome

We analyzed the relationship between QRMs and treatment outcome of quinolone-based therapy. Of the 128 patients who were treated with quinolone-based therapy, 39 were not evaluated with a TOC. Finally, we compared variables between the treatment failure group (*n* = 27) and treatment success group (*n* = 62) (**Table 3**). Except for 5 patients, sequential therapy with minocycline and sitafloxacin was administered. No differences were observed in terms of sex (*p* = 0.767) or age (*p* = 0.446). Regarding *parC* mutations, using non-S83I as a reference, the S83I mutation was associated with an increased treatment failure rate with a marginally significant difference (37.3%, 22/59 vs. 16.7%, 5/30, respectively, *p* = 0.054). For *gyrA* mutations, with WT as a reference, the mutant type was significantly associated with increased treatment failure (52.4%, 11/21 vs. 23.5%, 16/68, respectively, *p* = 0.016). In the combinations of *parC* and *gyrA*, the treatment failure rate was higher in cases with *parC* S83I and *gyrA* mutations than in those with *parC* S83I and WT *gyrA*. With non-S83I:WT as a reference, the S83I:mutant combination was significantly associated with treatment failure (50.0%, 10/20 vs. 13.8%, 4/29, respectively, *p* = 0.010). There was no difference for the S83I:WT combination (30.8%, 12/39 vs. 13.8%, 4/29, respectively, *p* = 0.150).

**Table 3.** Comparison of *parC* and *gyrA* mutations between the treatment failure and success cases.

|  |  | Treatment outcome |  |  |
| --- | --- | --- | --- | --- |
| Variable |  | Failure (*n* = 27) | Success (*n* = 62) | *p*-value |
| Sex, *n* (%) | Male | 23/72 (31.9) | 49/72 (68.1) | Reference |
|  | Female | 4/17 (23.5) | 13/17 (76.5) | 0.767 |
| Age, years, median [interquartile range] |  | 29 [24–35] | 30 [27–35] | 0.446 |
| *parC*, *n* (%) |  |  |  |  |
|  | Non-S83I | 5/30 (16.7) | 25/30 (83.3) | Reference |
|  | WT | 2/6 (33.3) | 4/6 (66.7) | - |
|  | S83N | 1/14 (7.1) | 13/14 (92.9) | - |
|  | D82N | 0/– | 1/1 (100) | - |
|  | D87Y | 0/– | 4/4 (100) | - |
|  | D87H | 0/– | 1/1 (100) | - |
|  | S83R | 1/3 (33.3) | 2/3 (66.7) | - |
|  | D87N | 1/1 (100) | 0/– (0) | - |
|  | S83I | 22/59 (37.3) | 37/59 (62.7) | 0.054 |
| *gyrA*, *n* (%) |  |  |  |  |
|  | WT | 16/68 (23.5) | 52/68 (76.5) | Reference |
|  | Mutant | 11/21 (52.4) | 10/21 (47.6) | 0.016* |
|  | M95I | 6/15 (40.0) | 9/15 (60.0) | - |
|  | D99N | 3/3 (100) | 0/– (0) | - |
|  | M95V | 1/2 (50.0) | 1/2 (50.0) | - |
|  | G93C | 1/1 (100) | 0/– (0) | - |
| *parC:gyrA*, *n* (%) |  |  |  |  |
|  | non-S83I:WT | 4/29 (13.8) | 25/29 (86.2) | Reference |
|  | non-S83I:mutant | 1/1 (100) | 0/- | - |
|  | S83I:WT | 12/39 (30.8) | 27/39 (69.2) | 0.150 |
|  | S83I:mutant | 10/20 (50.0) | 10/20 (50.0) | 0.010* |

WT, wild type. Asterisk indicates a significant difference.

## Discussion

In this study, we demonstrated a high prevalence of MRMs and QRMs in MG collected in Tokyo, Japan, during the 2023–2025 period. Notably, MRMs were detected in 94.4% of the samples. The most common QRM was the *parC* S83I mutation, present in 65.5% of samples, while dual-QRMs (i.e., mutations in *parC* and *gyrA*) were observed in 22.5% of cases. Phylogenetic analysis based on the *mgpB*-*MG309* region further revealed that strains harboring MRMs and *parC* S83I, indicative of dual-class resistance, clustered within specific *mgpB* alleles.

Deguchi et al. analyzed 627 clinical samples collected between 2013 and 2017 and reported a marked rise in resistance-associated mutations during this period in Japan. Specifically, the prevalence of MRMs increased from approximately 40% to 70%, while the *parC* S83I mutation rate rose from 20% to 30%, and *gyrA* mutations increased from approximately 2% to 10% (11). More recently, Ando et al. examined 180 clinical samples collected from MSM between 2019 and 2022 and found that the prevalence of resistance-associated mutations continued to rise in Tokyo. Their study reported an MRM prevalence of 89.4%, *parC* S83I at 71.4%, and *gyrA* mutations at 22.5% (9). Our findings closely align with those of Ando et al., indicating that resistance rates have continued to increase in the 2020s compared with the 2010s. Although data on sexual orientation were not available in our study, it was conducted at clinics primarily serving patients with internal medicine and sexually transmitted infections. Considering this setting, as well as the absence of anal samples collected during the study period, it is likely that the majority of participants were heterosexual. These findings suggest that the increase in drug-resistant MG is not limited to the MSM population but is also occurring among heterosexual individuals.

Previous reports on MG strain diversity in Japan during the 2010s are limited. Kikuchi et al. analyzed 20 strains collected from men with urethritis in Sendai between 2011 and 2012, reporting MRMs in 25.0% of strains and *parC* S83I mutations in 15.0% (19). In that study, *mgpB* allele 7 (40.0%) was the most frequent, corresponding to Cluster B, followed by alleles 21 (15.0%, Cluster A) and 14 (15.0%, Cluster B). In contrast, although allele 7 remained commonly detected in the present study, there was also a high prevalence of *mgpB* alleles 161, 184, 79, and 140, which harbored a high frequency of dual-class resistance mutations—including MRMs and *parC* S83I—indicating a shift in the predominant circulating strains over the past decade. This shift may reflect the expansion of multiple clonal complexes of dual-class drug-resistant strains in Japan, potentially driven by antibiotic selective pressure. A similar phenomenon has been described in France, where Sabine et al. reported the emergence of *mgpB* type ST159 (closest to allele 140 in this study), which harbored dual-class drug-resistant *23S rRNA* 2059T and *parC* S83I in 2021–2022 (25). Unlike *parC* S83I, *gyrA* mutations were found sporadically and were not cluster-specific, suggesting that dual-QRMs may be emerging independently across different genetic backgrounds. These findings raise concerns about the spread of genetically diverse, dual-class drug-resistant clonal complexes, emphasizing the need for cross-regional molecular surveillance.

In the treatment of MG, sequential therapy using tetracycline-macrolide or tetracycline-fluoroquinolone combinations is gaining attention (26). The cure rate for MG with doxycycline monotherapy is 30%–40% (27), but it has been shown that doxycycline treatment can reduce the bacterial load of MG (26). In Melbourne, Australia, during 2016–2017, a treatment strategy involving doxycycline pretreatment followed by the administration of azithromycin or sitafloxacin/moxifloxacin based on MRMs achieved a cure rate of over 92% (26). This approach was applied to a population in which two-thirds of cases harbored MRMs, and 20% of those with MRMs also had QRMs (7, 8, 26). In contrast, the overall cure rate in the present study was lower, at 69.7% (62 out of 89 patients), compared with the above-mentioned study. The high prevalence of *parC* S83I and *gyrA* mutations in our study population is likely to have contributed to the low cure rate. Several studies have demonstrated that the *parC* S83I mutation is associated with an increased risk of quinolone treatment failure, with failure rates ranging from 21.1% to 60% (6, 9, 22). Furthermore, when *parC* S83I and *gyrA* mutations are present, the treatment failure rate rises even further, reaching 58.3% to 81.2% (6, 9, 22). Our findings are consistent with previous studies, in which the treatment failure rate was 37.3% for the *parC* S83I mutation overall, but rose to 50% in the presence of a *gyrA* mutation. In Japan, *parC* S83I has become prevalent, resulting in a high treatment failure rate. However, with an increase in the prevalence of *gyrA* mutant strains, it is anticipated that the treatment failure rate will rise further. Promoting appropriate antimicrobial use and implementing interventions to prevent the spread of strains with *parC* S83I and *gyrA* mutations and maintain treatment success rates are strongly warranted.

With the increasing rate of treatment failure, refractory cases that do not respond to multiple courses of antibiotic therapy have been on the rise. A third-line treatment regimen has yet to be established; however, several options for refractory cases have been recommended, including pristinamycin and long-term minocycline and doxycycline (27). In this study, the majority of patients were cured through the re-administration of sequential therapy with minocycline and sitafloxacin and prolonged minocycline. Therefore, these regimens may be valuable options when initial therapy fails. However, up to seven courses were required for refractory MG infection in this study. The exploration of effective third-line treatments should be a priority.

The major limitation of this study is that sample collection was conducted only at clinics in Tokyo. Given that previous studies have identified geographical differences in antimicrobial resistance rates even in the same country (11), our findings may not fully reflect the nationwide distribution of circulating genotypes in Japan. Another limitation is that the dataset contained a considerable amount of missing TOC results, which may have affected the evaluation of treatment failure rates. Further epidemiological studies involving a broader range of regions are warranted to better understand the national trends in resistance and strain distribution.

In conclusion, we found a high prevalence of drug resistance-associated mutations in MG in Tokyo, Japan. Notably, potential dual-class drug-resistant clonal complexes with MRMs and the *parC* S83I mutation—particularly those harboring *mgpB* alleles 79, 140, 161, and 184—appear to be emerging. To maintain treatment success rates, it is essential to promote the appropriate use of antibiotics and implement ongoing surveillance to prevent a further increase in drug resistance-associated mutations in MG.

## Data Availability

All data produced in the present study are available upon reasonable request to the authors

## Consent for publication

Not applicable.

## Availability of data and materials

The datasets analyzed during the present study are available from the corresponding author on reasonable request.

## Acknowledgements

We thank the staff of KARADA Internal Medicine Clinics.

## Authors’ contributions

KI, study conceptualisation; RO, KT, and HM investigation; AS, MT, and NM, data collection and curation; RO, performing formal analysis; RO and KI, manuscript drafting; MT, KT, HM, and NM, manuscript revision; TM, study supervision; KI and TM, Funding acquisition. All authors have read and approved the final manuscript.

## Supplementary materials

**Supplementary Table S1**. Primers used in this study.

**Supplementary Table S2**. Twenty-three patients who achieved microbiological cure after initial treatment failure.

**Supplementary Table S3**. Frequency of dual-class drug resistance-associated mutations in 142 clinical samples.

**Supplementary Table S4**. Results of *mgpB*-*MG309* typing in this study.

## References

1. Gnanadurai R, Fifer H. 2020. Mycoplasma genitalium: A Review. Microbiology 166:21–29.

2. Ito S, Hanaoka N, Shimuta K, Seike K, Tsuchiya T, Yasuda M, Yokoi S, Nakano M, Ohnishi M, Deguchi T. 2016. Male non-gonococcal urethritis: From microbiological etiologies to demographic and clinical features. Int J Urol 23:325–31.

3. Chua TP, Vodstrcil LA, Murray GL, Plummer EL, Jensen JS, Unemo M, Chow EPF, Low N, Whiley DM, Sweeney EL, Hocking JS, Danielewski JA, Garland SM, Fairley CK, Zhang L, Bradshaw CS, Machalek DA. 2025. Evolving patterns of macrolide and fluoroquinolone resistance in Mycoplasma genitalium: an updated systematic review and meta-analysis. Lancet Microbe doi:10.1016/j.lanmic.2024.101047:101047.

4. Jensen JS, Bradshaw CS, Tabrizi SN, Fairley CK, Hamasuna R. 2008. Azithromycin treatment failure in Mycoplasma genitalium-positive patients with nongonococcal urethritis is associated with induced macrolide resistance. Clin Infect Dis 47:1546–53.

5. Hamasuna R, Le PT, Kutsuna S, Furubayashi K, Matsumoto M, Ohmagari N, Fujimoto N, Matsumoto T, Jensen JS. 2018. Mutations in ParC and GyrA of moxifloxacin-resistant and susceptible Mycoplasma genitalium strains. PLoS One 13:e0198355.

6. Murray GL, Plummer EL, Bodiyabadu K, Vodstrcil LA, Huaman JL, Danielewski JA, Chua TP, Machalek DA, Garland S, Doyle M, Sweeney EL, Whiley DM, Bradshaw CS. 2023. gyrA Mutations in Mycoplasma genitalium and Their Contribution to Moxifloxacin Failure: Time for the Next Generation of Resistance-Guided Therapy. Clinical Infectious Diseases 76:2187–2195.

7. Murray GL, Bradshaw CS, Bissessor M, Danielewski J, Garland SM, Jensen JS, Fairley CK, Tabrizi SN. 2017. Increasing Macrolide and Fluoroquinolone Resistance in Mycoplasma genitalium. Emerg Infect Dis 23:809–812.

8. Couldwell DL, Tagg KA, Jeoffreys NJ, Gilbert GL. 2013. Failure of moxifloxacin treatment in Mycoplasma genitalium infections due to macrolide and fluoroquinolone resistance. Int J STD AIDS 24:822–8.

9. Ando N, Mizushima D, Takano M, Mitobe M, Kobayashi K, Kubota H, Miyake H, Suzuki J, Sadamasu K, Aoki T, Watanabe K, Uemura H, Yanagawa Y, Gatanaga H, Oka S. 2023. Effectiveness of sitafloxacin monotherapy for quinolone-resistant rectal and urogenital Mycoplasma genitalium infections: a prospective cohort study. J Antimicrob Chemother 78:2070–2079.

10. Ando N, Mizushima D, Takano M, Mitobe M, Miyake H, Yokoyama K, Sadamasu K, Aoki T, Watanabe K, Uemura H, Yanagawa Y, Gatanaga H, Oka S. 2021. High prevalence of circulating dual-class resistant Mycoplasma genitalium in asymptomatic MSM in Tokyo, Japan. JAC Antimicrob Resist 3:dlab091.

11. Deguchi T, Ito S, Yasuda M, Sato Y, Uchida C, Sawamura M, Manda K, Takanashi M, Kiyota H. 2018. Surveillance of the prevalence of macrolide and/or fluoroquinolone resistance-associated mutations in Mycoplasma genitalium in Japan. J Infect Chemother 24:861–867.

12. Hjorth SV, Björnelius E, Lidbrink P, Falk L, Dohn B, Berthelsen L, Ma L, Martin DH, Jensen JS. 2006. Sequence-based typing of Mycoplasma genitalium reveals sexual transmission. J Clin Microbiol 44:2078–83.

13. Ma L, Martin DH. 2004. Single-Nucleotide Polymorphisms in the rRNA Operon and Variable Numbers of Tandem Repeats in the Lipoprotein Gene among <i>Mycoplasma genitalium</i> Strains from Clinical Specimens. Journal of Clinical Microbiology 42:4876–4878.

14. Dumke R. 2022. Molecular Tools for Typing Mycoplasma pneumoniae and Mycoplasma genitalium. Front Microbiol 13:904494.

15. Lara I, Hernandez-Ruiz V, Fernández-Huerta M, Rodriguez-Grande J, Arnaiz De Las Revillas F, Rodriguez-Lozano J, Calvo-Montes J, Ocampo-Sosa A, Fariñas MC, Roiz Mesones MP, Garcia-Fernandez S, Moure Z. 2025. Mycoplasma genitalium and antimicrobial resistance among the general female and male population in northern Spain. Sex Transm Infect doi:10.1136/sextrans-2024-056374.

16. Sweeney EL, Tickner J, Bletchly C, Nimmo GR, Whiley DM. 2020. Genotyping of Mycoplasma genitalium Suggests De Novo Acquisition of Antimicrobial Resistance in Queensland, Australia. J Clin Microbiol 58.

17. Pond MJ, Nori AV, Witney AA, Lopeman RC, Butcher PD, Sadiq ST. 2014. High prevalence of antibiotic-resistant Mycoplasma genitalium in nongonococcal urethritis: the need for routine testing and the inadequacy of current treatment options. Clin Infect Dis 58:631–7.

18. Ito S, Shimada Y, Yamaguchi Y, Yasuda M, Yokoi S, Ito S, Nakano M, Ishiko H, Deguchi T. 2011. Selection of Mycoplasma genitalium strains harbouring macrolide resistance-associated 23S rRNA mutations by treatment with a single 1 g dose of azithromycin. Sex Transm Infect 87:412–4.

19. Kikuchi M, Ito S, Yasuda M, Tsuchiya T, Hatazaki K, Takanashi M, Ezaki T, Deguchi T. 2014. Remarkable increase in fluoroquinolone-resistant Mycoplasma genitalium in Japan. J Antimicrob Chemother 69:2376–82.

20. Jensen JS, Björnelius E, Dohn B, Lidbrink P. 2004. Use of TaqMan 5’ nuclease real-time PCR for quantitative detection of Mycoplasma genitalium DNA in males with and without urethritis who were attendees at a sexually transmitted disease clinic. J Clin Microbiol 42:683–92.

21. Getman D, Jiang A, O’Donnell M, Cohen S. 2016. Mycoplasma genitalium Prevalence, Coinfection, and Macrolide Antibiotic Resistance Frequency in a Multicenter Clinical Study Cohort in the United States. J Clin Microbiol 54:2278–83.

22. Murray GL, Bodiyabadu K, Danielewski J, Garland SM, Machalek DA, Fairley CK, Jensen JS, Williamson DA, Tan LY, Mokany E, Durukan D, Bradshaw CS. 2020. Moxifloxacin and Sitafloxacin Treatment Failure in Mycoplasma genitalium Infection: Association with parC Mutation G248T (S83I) and Concurrent gyrA Mutations. J Infect Dis 221:1017–1024.

23. Jolley KA, Bray JE, Maiden MCJ. 2018. Open-access bacterial population genomics: BIGSdb software, the PubMLST.org website and their applications. Wellcome Open Res 3:124.

24. Nguyen L-T, Schmidt HA, von Haeseler A, Minh BQ. 2014. IQ-TREE: A Fast and Effective Stochastic Algorithm for Estimating Maximum-Likelihood Phylogenies. Molecular Biology and Evolution 32:268–274.

25. Pereyre S, Laurier-Nadalié C, Balcon C, Hénin N, Dolzy A, Gardette M, Guiraud J, Bébéar C. 2025. Spread of Dual-Resistant Mycoplasma genitalium Clone among Men, France, 2021-2022. Emerg Infect Dis 31:854–858.

26. Read TRH, Fairley CK, Murray GL, Jensen JS, Danielewski J, Worthington K, Doyle M, Mokany E, Tan L, Chow EPF, Garland SM, Bradshaw CS. 2019. Outcomes of Resistance-guided Sequential Treatment of Mycoplasma genitalium Infections: A Prospective Evaluation. Clin Infect Dis 68:554–560.

27. Jensen JS, Cusini M, Gomberg M, Moi H, Wilson J, Unemo M. 2022. 2021 European guideline on the management of Mycoplasma genitalium infections. J Eur Acad Dermatol Venereol 36:641–650.

